# Young males in crisis: Pathways, usage and acceptability of an online messenger based psychosocial counselling service

**DOI:** 10.1101/2025.11.04.25339230

**Authors:** Juliane Hug, Elisabeth Kohls, Konrad Jakob Endres, Melanie Eckert, Richard Wundrack, Shadi Saee, Juliane Pougin, Aneliana da Silva Prado, Christine Rummel-Kluge

## Abstract

**Introduction:** Boys and young men face an elevated risk of mental health problems and suicidality, yet they remain less likely than their female peers to seek professional help. Online counselling services such as *krisenchat* offer low-threshold support and may help reduce gender-specific barriers, but little is known about how young men use these services.

**Objective:** This study investigates male *krisenchat* users in comparison to other users, focusing on demographics, utilization patterns, satisfaction, chat topics, and barriers to help-seeking behavior, in order to generate insights for improving mental health support for young men.

**Methods:** Anonymized data were obtained from *n* = 29,387 *krisenchat* users between January and December 2023. After data cleaning, the final sample comprised of *N* = 9,584 participants. Demographic information, utilization behavior, suicidality, and use of professional help services were documented by counsellors, while user satisfaction, recommendation rates, and emotional distress were assessed through voluntary surveys following consultation.

**Results:** Young males accounted for 19.9% of *krisenchat* users, were on average older than female users and were less likely to have been in prior treatment. Male users sent fewer messages, accessed the service during late-night hours more often than females, and tended to find the service via search engines rather than institutional or social media channels. Compared to female users, they were less likely to disclose self-harm, family problems, or sexual violence, but more likely to bring up sexuality and LGBTQIA+ topics. Importantly, no gender difference was found for suicidality. Despite differences in some utilization patterns, acceptability outcomes — including reductions in distress, satisfaction, and likelihood of recommending the service — were comparable across genders, suggesting equivalent counselling benefits once engaged.

**Conclusions:** Digital crisis services like *krisenchat* hold potential for reducing gender disparities in mental health support. However, targeted strategies to improve visibility, adapt communication styles, and strengthen follow-up pathways are essential to increase engagement and sustained help-seeking among young men.

**Study Registration:** DRKS00026671

## 1. Introduction

The mental health of boys and young men is a challenging issue worldwide. Despite having a similar prevalence of mental disorders, boys and young men participate in health interventions far less frequently (1). This is concerning, as they represent a particular risk group for mental health problems, as they not only show higher rates of suicidal behaviour (2), conduct disorder, substance use, and interpersonal violence (3) but also significantly underutilize professional mental health services (4).

Recent statistics highlight the severity of this issue in Germany, where the overall suicide rate was 12.2 per 100,000 in 2023, with a marked gender gap: 17.9 per 100,000 for men versus 6.6 per 100,000 for women (5). Within younger age groups, statistics from 2023 show that among 15-to 20-year-olds, there were 173 suicides, of which 123 (71.1%) were male and 50 (28.9%) female (6). A large study of adolescents and young adults in Germany (age 14-21, *N* = 1,180) found that 10.7% reported lifetime suicidal ideation, 5.0% a suicidal plan, and 3.4% a suicide attempt (7). Although additional research on suicidality in young males is necessary, findings from a cross-sectional epidemiological study involving over 495 male participants aged 14 to 21 indicate a lifetime prevalence of suicidal ideation at 8.7% and suicide attempts at 2.3% within this population (7).

A contributing factor to this elevated risk is reduced help-seeking behavior. A large systematic review involving over 96,297 participants examined help-seeking behaviour among young adults (aged 18–24) and found that young women are more than twice as likely to seek help from mental health services compared to young men (8). Qualitative findings further indicate that young men face a range of barriers to seeking professional help such as fear of psychiatric medication, the influence of traditional masculine norms, and the concern among young gay men about potential homophobic responses from mental health professionals (4).

It is hypothesized that self-stigmatization and conformity to masculine social norms (CMN) in young men can hinder help-seeking behaviour and may increase the risk of suicide if the underlying psychiatric illness remains untreated (9). Leaving mental illnesses untreated is especially concerning, as psychiatric disorders such as depression and psychosis are leading causes of suicide worldwide (10).

While young men are less likely than young women to use traditional mental health services, online mental health services may help to close this gap, as they are easily accessible and offer low-threshold support (11). While there is a growing body of literature on online chat counselling for mental health (12–17), research focusing especially on young men is still scarce.

Strengthening the evidence base for male usage, preferences and feasibility of online counselling programs essential. *krisenchat*, launched in Germany in 2020, has proven feasible and acceptable, with most users being young and female (13). The service has shown benefits for acute crises such as suicidality (18), and a high proportion (48.6%) of users follow recommendations for further help (15). However, little is known about how young males use such services, whether their needs and help-seeking differ, or how effective online interventions are in overcoming gender-specific barriers. Addressing this gap is crucial, as young men remain at higher risk of suicide yet are less likely to engage with traditional mental health services.

Against this background, the present study aims to describe and investigate users of *krisenchat* who identified as male, comparing them to other users in terms of demographic characteristics, utilization behaviour, satisfaction, chat topics, and help-seeking patterns (including barriers). By addressing these questions, this study seeks to generate insights into the characteristics and needs of male users, and to evaluate strategies for improving their engagement with online services. Ultimately, the findings aim to contribute to a better understanding of help-seeking behaviour among young men and inform approaches to increase mental health service uptake in this population.

## 2. Materials and Methods

### 2.1 Description of the krisenchat counselling service

*Krisenchat* (www.krisenchat.de) is a service provided by both volunteer and on-staff counsellors with academic backgrounds in psychology or social work that assists children and young adults in acute crises. It has been offered in a free, confidential, and around the clock chat routine since May 2020. For more information on the service, please see previous works (13, 15, 16, 19–21).

### 2.2. Participants and Procedure

Anonymized data from all users of *krisenchat* were extracted from the operational database between January 1, 2023 and December 31, 2023. Data encompasses metadata, data collected by counsellors during chat sessions, users’ responses to a feedback survey shortly after the session, and a follow-up survey after four weeks. Procedure on feedback and follow-up survey is presented in detail in (15).

Informed consent was inquired via an opt-in function prior to survey participation. The Ethics Committee of the Medical Faculty at the University of Leipzig granted ethical approval in April 2025 (file reference: 372/21-ek).

Data of *N* = 29,387 *krisenchat* users were collected during the above mentioned period. Exclusion criteria were the following: users without the serious intention of getting a counselling session or without an ongoing crisis (*n* = 20). Data from the Ukrainian platform (*n* = 8,999). Participants excluded due to missing data (*n* = 9,631), and for being older than 25 years (*n* = 11) – totalizing *n* = 9,644 excluded cases in this step. Finally, participants who were flagged as suspected cases of child welfare endangerment were also excluded (*n* = 1,144).

Hence, data of *N* = 9,584 users were analyzed (see Figure 1). From this final sample, *n* = 2,836 (29.6%) completed the feedback survey, and *n* = 1,191 (12.4%) completed the follow-up survey.

**Figure 1.**
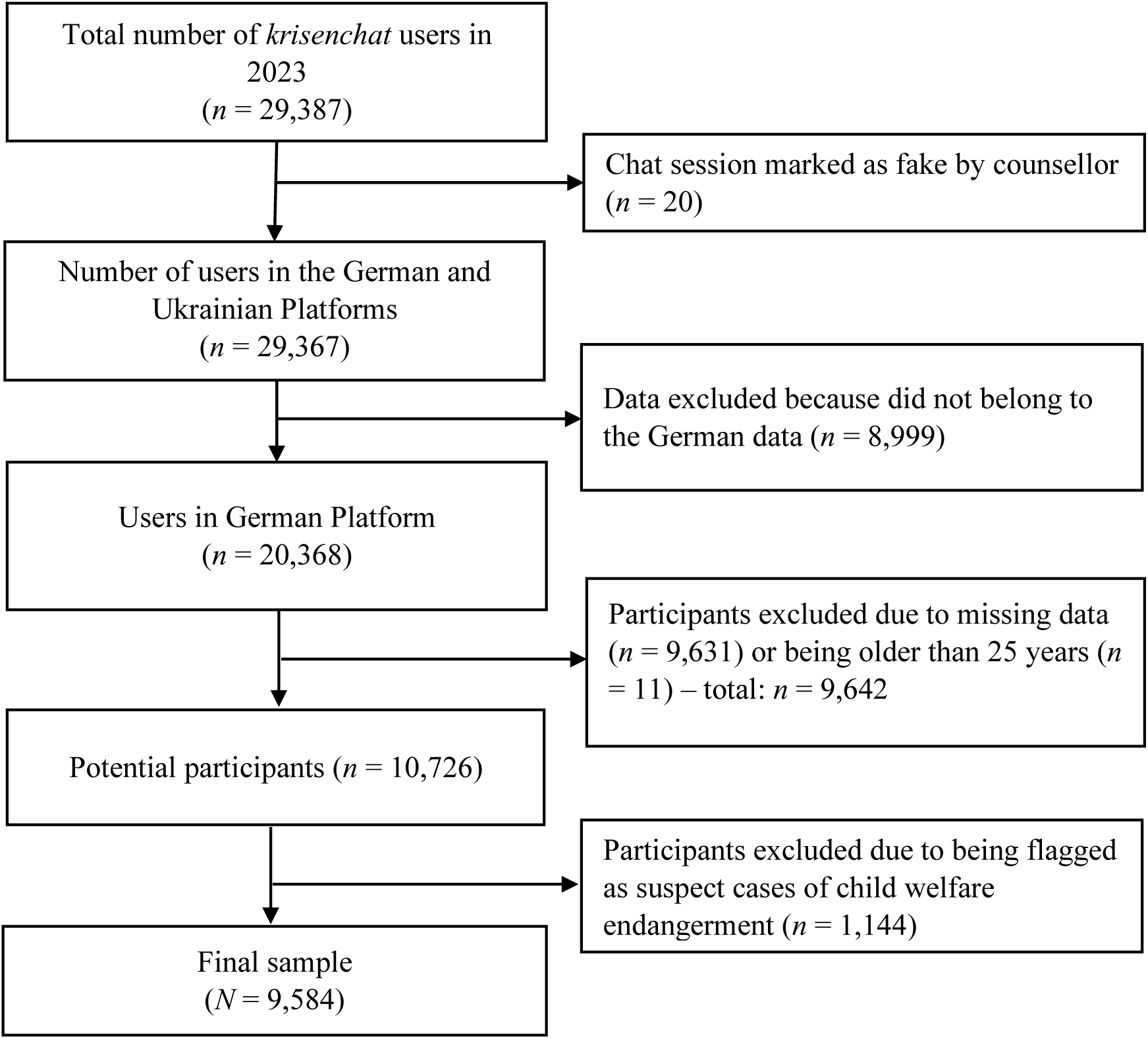
Flowchart of data cleaning process.

### 2.3. Measures

#### Demographic Information

Data on users’ age, gender, and prior or current use of professional help providers were collected and noted by the counsellors during the chat, if users disclosed them. Additionally, general ‘chats-at-risk’ – namely sexual violence and suicidal behavior were identified by the counsellors.

#### Utilization behavior

Metadata and data collected by counsellors during chat sessions were used to assess utilization behavior, including: total number of messages sent by the user and by the counsellor during the entire consultation period, time of contacting the service, and matters addressed by users classified by counsellors using chat topics.

Additionally, users were asked about how they learned about service (e.g., social media, school, recommendation) in the feedback survey.

#### Acceptability and help seeking behavior

In the feedback survey, participants were asked to indicate the perceived helpfulness of the service and the likelihood of service recommendation. Perceived helpfulness was assessed using a four-point Likert scale that further was computed by dichotomizing the scale into “yes” (2 = “rather yes,” 3 = “yes”) and “no” (0 = “no,” 1 = “rather no”) (21). Participants were asked to indicate how likely they were to recommend the service on a Likert scale ranging from 0% = “not at all” to 100 % = “definitely,” using an adapted version of the Net Promoter Score (NPS; (22). The likelihood of service recommendation was calculated by converting the rate into a binary variable, indicated by a score of 6 or higher (18, 21).

The brief Screening Tool for Psychological Distress (STOP-D; (23)) was used to evaluate potential differences in users’ distress before and shortly after the chat. The STOP-D consists of five items (depressive symptoms, anxiety, stress, anger, and insufficient social support) assessed on a 10-point Likert scale (0 = “not at all” to 9 = “very much”). A sum score was calculated, resulting in STOP-D pre-and post-values as well as a change score for each participant.

In the follow-up survey, users were asked if they contact the person or professional help services they were referred to (e.g., specialized professional counselling service, school psychologist or school social worker, social services, local youth center, psychotherapist or social psychiatric service, general practitioner, calling an ambulance, parents, or another trusted adult person). Users who indicated having followed the recommendation they received, were considered as displaying further help-seeking behavior.

Users who reported not seeking further help, regardless of having received a recommendation, had their barriers evaluated. Based on previous work (15, 24), barriers were grouped as follows (see Table 1):

**Table 1.** Barriers to seeking further help among *krisenchat* users.

| Barriers | Description |
| --- | --- |
| **Stigma** | Scared of being placed in a psychiatric unit, or not feeling confident enough to open up to a stranger. |
| **Lack of literacy** | Fear of not being taken seriously, believing that it is one's own fault, or not believing that the referral would help. |
| **Structural** | The service is too far or hard to reach, or there was no time to contact it yet. |
| **Autonomy** | No longer needing help, feeling better, or feeling able to deal with the issue independently |
| **Family beliefs** | Fear that a parent would find out. |
| **Other reasons** | Other reasons not specified above. |

The General Self-efficacy Short Scale (German: *Allgemeine Selbstwirksamkeit Kurzskala* [ASKU]) (25) was used to assess users’ general self-efficacy beliefs. The scale consists of three items, each representing a single statement (e.g., “I am able to solve most problems on my own”), rated on a 5-point Likert scale ranging from 1 = “does not apply at all” to 5 = “fully applies.” Higher mean scores indicate higher levels of self-efficacy.

### 2.4. Statistical Analysis

All statistical analyses were performed using IBM SPSS Statistics version 29.0. A two-tailed α = 0.05 was applied to statistical testing.

First, descriptive statistics were performed for demographic information. Second, diverse, female and male chatters were analyzed separately. We used chi-square tests to estimate subgroup differences. Following, when significant effects were found using chi-square tests, we applied *z*-tests to compare proportions.

One-way ANOVAS were used to assess differences between the gender subgroups (diverse, female, and male) on age, likelihood of recommendation, user satisfaction, STOP-D pre-and post-chat session, mean of total messages sent by chatter and by counsellor. The assumption of homogeneity of variance was assessed using Levene’s test. Considering the heterogeneity of variance, Welch’s correction was requested and post-hoc evaluation using the Games-Howell technique was applied instead of Tukey technique (26).

Subsequently, to compare emotional distress (STOP-D) before and after consultation, we used a paired sample *t*-test for each gender.

We applied a bootstrapping procedure (1,000 resamplings; 95% bias-corrected and accelerated confidence intervals [CI BCa]) to correct for group size differences and deviations from a normal distribution, and to present 95% confidence intervals for the means (27). We also used Bonferroni correction to adjust for multiple testing when applicable. Effect sizes for chi-squared tests were estimated using the ϕ coefficient; when the contingency table was larger than 2 × 2, Cramér’s *V* (ϕc) was used with ϕ, ϕc = 0.10 indicating a small effect, ϕ, ϕc = 0.30 indicating a medium effect, and ϕ or ϕc = 0.50 a large effect. The effect size for *t*-tests was interpreted as small when *d* < 0.20, medium when *d* < 0.50 and large when *d* > 0.50 (28). To estimate the effect size for the ANOVAs, η2 was interpreted as small when η2 = 0.001, as medium when η2 = 0.06, and as large when η2 = 0.14 (28).

## 3. Results

### 3.1. Demographic Information

Of the *N* = 9,582 users included in the analysis, *n* = 7,261 (75.8%) were female, *n* = 1,904 (19.9%) were male, and *n* = 296 (3.1%) were diverse. Gender information was missing for *n* = 121 (1.3%) users. The age of the total sample ranged from 9 to 25 years (*M* = 17.36, *SD* = 3.59). Among the total sample, *n* = 528 (5.5%) reported sexual violence, *n* = 398 (4.2%) presented suicidal behavior, *n* = 893 (9.3%) were already receiving professional help in the support system, and *n* = 2,184 (22.8%) received a recommendation to seek further support.

The recommendation rate of the service was high, with 99.1% (*n* = 2,503) of participants recommending the service. Most participants (*n* = 2,264, 89.6%) indicated the service helped them well or very well.

Detailed demographic information for each gender subgroup (male, female, and diverse) is presented in Table 2. Male users (*M* = 18.12, *SD* = 3.42) were older than female (*M* = 17.17, *SD* = 3.62) and diverse (*M* = 17.01, *SD* = 3.18) users and less likely to report sexual violence (*n* = 56, 2.9%) compared to female (*n* = 451, 6.2%) users, but did not statistically differ from diverse users (*n* = 15, 5.1%) in this regard. They (*n* = 83, 4.4%) were also less likely to report suicidal behavior compared to diverse users (*n* = 23, 7.8%) but did not statistically differ from female users (*n* = 286, 3.9%). Male users (*n* = 317, 16.6%) were less likely to be receiving current treatment or intervention compared to both female (*n* = 1,828, 25.2%) and diverse users (*n* = 93, 31.4%).

**Table 2.** Group comparison between male, female and diverse users (*N* = 9,461).

| Variable, $n$ (%) | Male<br>($n = 1,904$) | 95%-CI<br>BCa | Female<br>($n = 7,261$) | 95%-CI<br>BCa | Diverse<br>($n = 296$) | 95%-CI<br>BCa | Test | $p$ | Effect<br>size |
| --- | --- | --- | --- | --- | --- | --- | --- | --- | --- |
| | $n$ (%) | | $n$ (%) | | $n$ (%) | | $\chi^2$ | | $\phi_c$ |
| Sexual violence | 56 (2.9)<sup>a</sup> | NA | 451 (6.2)<sup>b</sup> | NA | 15 (5.1)<sup>a,b</sup> | NA | $\chi^2(2, 9,461) = 31.062$ | < .001 | .057 |
| Suicidal behavior | 83 (4.4)<sup>a</sup> | NA | 286 (3.9)<sup>a</sup> | NA | 23 (7.8)<sup>b</sup> | NA | $\chi^2(2, 9,461) = 10.792$ | .005 | .034 |
| Prior use of professional help services | 130 (29.7)<sup>a</sup> | NA | 717 (34.9)<sup>a</sup> | NA | 34 (42.0)<sup>a</sup> | NA | $\chi^2(2, 2,573) = 6.603$ | .037 | .051 |
| Recommended to other support services | 437 (23.0)<sup>a</sup> | NA | 1,668 (23.0)<sup>a</sup> | NA | 56 (18.9)<sup>a</sup> | NA | $\chi^2(2, 9,461) = 2.668$ | .263 | .017 |
| Current treatment<br>or intervention | 317 (16.6)<sup>a</sup> | NA | 1,828<br>(25.2)<sup>b</sup> | NA | 93 (31.4)<sup>c</sup> | NA | $\chi^2(2, 9,461) = 70.923$ | < .001 | .087 |
| Further help-seeking | 61 (43.0)<sup>a</sup> | NA | 394 (48.9)<sup>a</sup> | NA | 12 (31.6)<sup>a</sup> | NA | $\chi^2(2, 985) = 5.708$ | .058 | .076 |
| | *M (SD)* | | *M (SD)* | | *M (SD)* | | *F*-test | | $\eta^2, \omega^2$ |
| Age, *M (SD)* | 18.12<br>(3.42)<sup>a</sup> | 17.97-18.29 | 17.17<br>(3.62)<sup>b</sup> | 17.09-17.25 | 17.01<br>(3.18)<sup>b</sup> | 16.66-17.36 | Welch's *F*(2,<br>770,867) = 58.558 | < .001 | .011 |
| Likelihood of<br>recommendation,<br>*M (SD)* | 83.55<br>(22.60)<sup>a</sup> | 81.51-85.43 | 85.93<br>(20.08)<sup>a</sup> | 85.09-86.72 | 85.19<br>(23.49)<sup>a</sup> | 79.03-90.92 | Welch's *F*(2,<br>186,326) = 2.340 | .135 | .001 |
| User satisfaction,<br>*M (SD)* | 2.33 (0.72)<sup>a</sup> | 2.26-2.39 | 2.32 (0.72)<sup>a</sup> | 2.29-2.35 | 2.27 (0.64)<sup>a</sup> | 2.11-2.43 | *F*(2, 2,496) = 0.189 | .828 | .000 |
| STOP-D pre | 6.31 (1.61)<sup>a</sup> | 6.17-6.47 | 6.44 (1.55)<sup>a</sup> | 6.37-6.51 | 6.60 (1.31)<sup>a</sup> | 6.29-6.87 | Welch's *F*(2,<br>207,059) = 1.909 | .151 | .001 |
| STOP-D post | 4.44 (2.20)<sup>a</sup> | 4.22-4.65 | 4.60 (2.23)<sup>a</sup> | 4.51-4.70 | 4.66 (1.98)<sup>a</sup> | 4.21-5.11 | *F*(2, 2,620) = 1.074 | .342 | .001 |
| Mean of total<br>messages sent by<br>chatter, *M (SD)* | 53.04<br>(76.27)<sup>a</sup> | 49.98-56.45 | 61.90<br>(111.24)<sup>b</sup> | 59.24-64.79 | 59.96<br>(83.92)<sup>a,b</sup> | 51.78-70.16 | Welch's *F*(2,<br>796,920) = 8.267 | < .001 | .001 |
| Mean of total<br>messages sent by<br>counsellor, *M*<br>(*SD*) | 36.17<br>(54.56)<sup>a</sup> | 34.02-38.86 | 42.23<br>(65.24)<sup>b</sup> | 40.87-43.66 | 43.96<br>(67.93)<sup>a,b</sup> | 37.81-52.97 | Welch's *F*(2,<br>760,365) = 8.886 | < .001 | .001 |
| Self-efficacy | 2.88 (0.94)<sup>a</sup> | 2.73-3.05 | 2.72 (0.90)<sup>a</sup> | 2.66-2.78 | 2.72 (0.83)<sup>a</sup> | 2.42-3.01 | *F*(2, 910) = 1.804 | .165 | .002 |

Notes. Calculation of % from valid cases. $\chi^2$ , Chi-Square-Test-Statistic; $\phi_c$ , Cramér's *V*; $\eta^2$ , Eta Square; $\omega^2$ , Omega Square. The different superscript letters <sup>a,b,c</sup> indicate significant differences between the gender subgroups (one-way ANOVAs) based on Bonferroni, Tukey, and Games-Howell post-hoc analysis.

There was no statistical difference between the three subgroups in prior use of professional services or in receiving a recommendation to seek further support.

### 3.2. Utilization behavior

The results indicated a statistically significant effect for the number of messages sent by chatters and counsellors. Male participants sent fewer messages (*M* = 53.04, *SD* =76.27) and received fewer messages (*M* = 36.17, *SD* = 54.56) compared to female users (respectively, *M* = 61.90, *SD* = 111.24; *M* = 42.23, *SD* =65.24), although the effect size was small. No significant difference was found between male and diverse users or between female and diverse users concerning the mean number of total messages users sent (*M* = 59.24, *SD* = 83.92) or received (*M* = 43.96, *SD* = 67.93).

More than one-third of all users (*n* = 3,487, 36.9%) contacted *krisenchat* for the first time between 8:00 PM and 12:00 AM. Approximately one-quarter (*n* = 2,305, 24.4%) reached out between 4:00 PM and 8:00 PM. A significant difference was found between the three groups in the time of first contact, χ^2^(10, 9,461) = 34.394, *p* <.001, ϕ_c_ =.043. Bonferroni-adjusted post hoc analyses showed that male users were more likely than female users to contact the service between 12:00 AM and 4:00 AM and between 4:00 AM and 8:00 AM, and less likely to contact between 8:00 PM and 12:00 AM. Diverse users did not differ significantly from either group. No significant differences were found across the other time ranges (see Table 3).

**Table 3.**
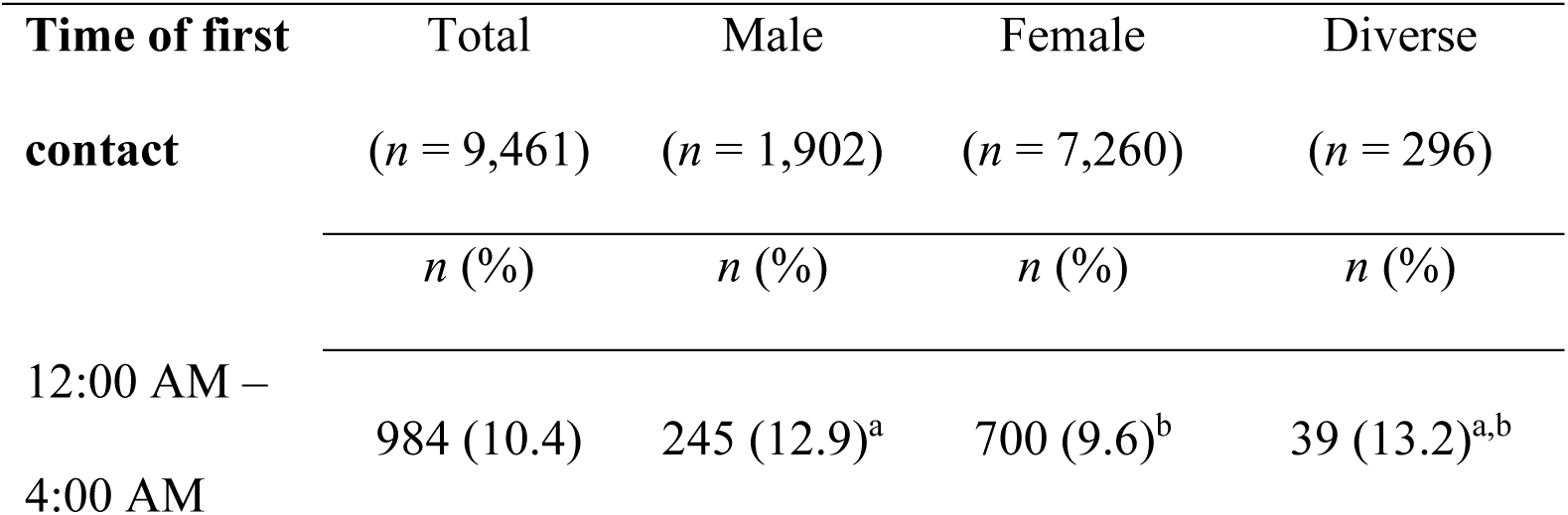
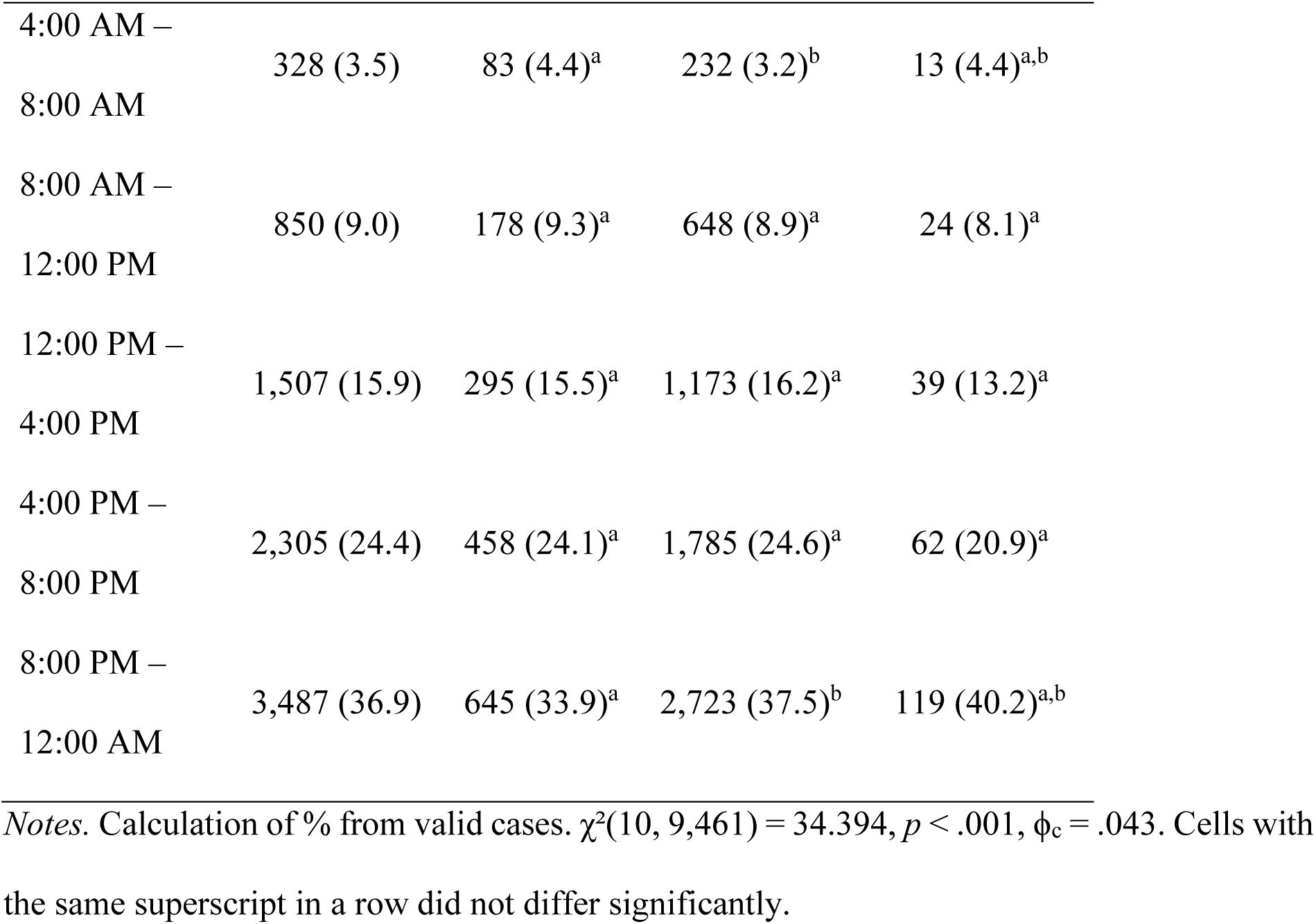
Group comparison between male, female, and diverse users regarding time of first contact to *krisenchat* (*N* = 9,461).

Among all users who answered the feedback survey (*n* = 2,836), most reported finding out about the service via own search on Google (*n* = 1,141, 45.7%), TikTok (*n* = 460, 18.4%), and friends and acquaintances (*n* = 439, 17.6%), see Table 4. There was a significant difference in sources of awareness across gender subgroups, χ^2^(16, 2,469) = 35.687, *p* =.003, ϕ_c_ =.085.

**Table 4.** Sources of awareness through which users first learned about the service, overall and stratified by gender identity (*n* = 2,836).

| Source of awareness | Total<br>($n = 2,836$) | Male<br>($n = 423$) | Female<br>($n = 1,970$) | Diverse<br>($n = 76$) |
| --- | --- | --- | --- | --- |
| | $n$ (%) | $n$ (%) | $n$ (%) | $n$ (%) |
| Own search (Google) | 1,141 (45.7) | 225 (53.2)<sup>a</sup> | 866 (44.0)<sup>b</sup> | 39 (51.3)<sup>a,b</sup> |
| TikTok | 460 (18.4) | 51 (12.1)<sup>a</sup> | 394 (20.0)<sup>b</sup> | 9 (11.8)<sup>a,b</sup> |
| Friends & acquaintances | 439 (17.6) | 77 (18.2)<sup>a</sup> | 340 (17.3)<sup>a</sup> | 16 (21.1)<sup>a</sup> |
| Instagram | 136 (5.4) | 20 (4.7)<sup>a</sup> | 114 (5.8)<sup>a</sup> | 2 (2.6)<sup>a</sup> |
| School or university | 114 (4.6) | 9 (2.1)<sup>a</sup> | 99 (5.0)<sup>b</sup> | 3 (3.9)<sup>a,b</sup> |
| YouTube | 105 (4.2) | 23 (5.4)<sup>a</sup> | 77 (3.9)<sup>a</sup> | 3 (3.9)<sup>a</sup> |
| Press (TV, radio, newspaper) | 32 (1.3) | 6 (1.4)<sup>a</sup> | 25 (1.3)<sup>a</sup> | 1 (1.3)<sup>a</sup> |
| Other | 70 (2.8) | 12 (2.9)<sup>a</sup> | 55 (2.8)<sup>a</sup> | 3 (3.9)<sup>a</sup> |

Note. Calculation of % from valid cases. $\chi^2(16, 2,469) = 35.687, p = .003, \phi_c = .085$ . Cells with expected counts $< 5$ may need to be excluded. When excluded “press” and “other”, then, $\chi^2(10, 2,367) = 31.981, p < .001, \phi_c = .082$ . Cells with the same superscript in a row did not differ significantly.

Bonferroni-adjusted post hoc analysis indicated that male users (*n* = 225, 53.2%) were more likely to discover the service via own search on Google compared to female users (*n* = 866, 44.0%); no significant difference was found between diverse users (*n* = 39, 51.3%) and both female and male users. Conversely, female users (*n* = 99, 5.0%; *n* = 394, 20.0%) were more likely to find about the service in school or university and on Tiktok compared to male users (*n* = 9, 2.1%; *n* = 51, 12.1%, respectively). No significant differences were found for diverse users (*n* = 3, 3.9%; *n* = 9; 11.8%) compared to both female and male users.

A comparison between male, female, and diverse users regarding chat topics indicated that male users were more likely to disclose emotional distress and sexuality compared to both female and diverse users. Conversely, they were less likely to report self-harm compared to both female and diverse users. No statistically significant difference was found between female and diverse users concerning self-harm. There was no significant difference between male and female users in reporting suicidality as a chat topic, while diverse users were more likely to report suicidality compared to both male and female users.

Male users were less likely to report family-related problems compared to female users, but not compared to diverse users. No statistically significant difference was found between female and diverse users regarding family-related problems.

Female users were more likely to report sexual violence compared to male users, but not compared to diverse users. No statistically significant difference between diverse and male users reporting sexual violence as a chat topic.

Diverse users were more likely to report LGBTQIA+ topics compared to both female and male users. Male users were more likely than female users to present LGBTQIA+ topics. For complete information about issues addressed by users as classified by counsellors using chat topics, see Table 5.

**Table 5.** Group comparison between male, female, and diverse users for chat topics and utilization behavior (*N* = 9,461).

| **Chat topics<sup>1,2</sup>** | Male | Female | Diverse | Test | $p$ | Effect size |
| --- | --- | --- | --- | --- | --- | --- |
| | ($n = 1,904$) | ($n = 7,261$) | ($n = 296$) | | | |
| | $n$ (%) | $n$ (%) | $n$ (%) | $\chi^2$ | | $\phi_c$ |
| **Mental disorders symptoms** | 626 (32.9)<sup>a</sup> | 2,700 (37.2)<sup>b</sup> | 112 (37.8)<sup>a,b</sup> | (2, 9,461) = 12.392 | .002 | .036 |
| Self-harm | 155 (8.1)<sup>a</sup> | 1,451 (20.0)<sup>b</sup> | 76 (25.7)<sup>a</sup> | (2, 9,461) = 157.767 | < .001 | .129 |
| Suicidality | 302 (15.9)<sup>a</sup> | 1,186 (16.3)<sup>a</sup> | 85 (28.7)<sup>b</sup> | (2, 9,461) = 32.463 | < .001 | .059 |
| **Family-topics** |  |  |  |  |  |  |
| Family-related problems | 220 (11.6)<sup>a</sup> | 1,196 (16.5)<sup>b</sup> | 43 (14.5)<sup>a,b</sup> | (2, 9,461) = 28.148 | < .001 | .055 |
| Acutely impaired parent-child relationship | 19 (1.0)<sup>a</sup> | 101 (1.4)<sup>a</sup> | 3 (1.0)<sup>a</sup> | (2, 9,461) = 2.012 | .366 | .015 |
| **Psychosocial distress** | 843 (44.3)<sup>a</sup> | 3,206 (44.2)<sup>a</sup> | 123 (41.6)<sup>a</sup> | (2, 9,461) = 0.810 | .667 | .009 |
| **Emotional distress** | 635 (33.4)<sup>a</sup> | 2,014 (27.7)<sup>b</sup> | 67 (22.6)<sup>b</sup> | (2, 9,461) = 28.731 | < .001 | .055 |
| **Violence** |  |  |  |  |  |  |
| Sexual violence | 43 (2.3)<sup>a</sup> | 350 (4.8)<sup>b</sup> | 10 (3.4)<sup>a,b</sup> | (2, 9,461) = 24.858 | < .001 | .051 |
| Family violence | 16 (0.8)<sup>a</sup> | 108 (1.5)<sup>a</sup> | 4 (1.4)<sup>a</sup> | (2, 9,461) = 4.7320 | .094 | .022 |
| Physical violence | 17 (0.9)<sup>a</sup> | 92 (1.3)<sup>a</sup> | 1 (1.2)<sup>a</sup> | (2, 9,461) = 3.647 | .161 | .020 |
| Partner violence | 10 (0.5)<sup>a</sup> | 107 (1.5)<sup>b</sup> | 3 (1.3)<sup>a,b</sup> | (2, 9,461) = 10.993 | .004 | .034 |
| Psychological violence | 19 (1.0)<sup>a</sup> | 138 (1.9)<sup>b</sup> | 1 (0.3)<sup>a,b</sup> | (2, 9,461) = 10.787 | .005 | .034 |
| **Education** | 36 (1.9)<sup>a</sup> | 160 (2.2)<sup>a</sup> | 5 (1.7)<sup>a</sup> | (2, 9,461) = 0.988 | .610 | .010 |
| **LGBTQIA+** | 104 (5.5)<sup>a</sup> | 80 (1.1)<sup>b</sup> | 46 (15.5)<sup>c</sup> | (2, 9,461) = 342.310 | < .001 | .190 |
| **Sexuality** | 67 (3.5)<sup>a</sup> | 78 (1.1)<sup>b</sup> | 2 (0.7)<sup>b</sup> | (2, 9,461) = 60.478 | < .001 | .080 |
| **COVID-19** | 3 (0.2)<sup>a,b</sup> | 8 (0.1)<sup>b</sup> | 2 (0.7)<sup>a</sup> | (2, 9,461) = 6.699 | .035 | .027 |
| **War** | 13 (0.7)<sup>a</sup> | 31 (0.4)<sup>a</sup> | -<sup>a</sup> | (2, 9,461) = 3.561 | .169 | .019 |

*Notes.* <sup>1</sup>Calculation of % from valid cases. <sup>2</sup>Multiple answers were possible. Cells with the same superscript in a row did not differ significantly.

### 3.4. Acceptability and help seeking behavior

There was no significant statistical difference between male, female, and diverse users in the likelihood of recommending the service to others, user satisfaction, pre-and post-chat distress levels, or general self-efficacy (see Table 2).

When examining each subgroup separately, all of them showed a significant decrease in distress levels after the chat compared to before.

Among male users, a paired pre-and post-chat comparison indicated significantly lower emotional distress after the session (*M* = 4.44, *SD* = 2.20, 95% CI BCa [4.23, 4.63]) compared to before (*M* = 6.31, *SD* = 1.61, 95% CI BCa [6.16, 6.45]), *t*(_450_) = 20.851, *p* <.001, Cohen’s *d* = 1.91.

Among female users, a paired pre-and post-chat comparison indicated significantly lower emotional distress after the session (*M* = 4.60, *SD* = 2.23, 95% CI BCa [4.50, 4.70]) compared to before (*M* = 6.44, *SD* = 1.55, 95% CI BCa [6.36, 6.51]), *t*(_2,087_) = 41.504, *p* <.001, Cohen’s *d* = 2.02.

Among diverse users, a paired pre-and post-chat comparison indicated significantly lower emotional distress after the session (*M* = 4.66, *SD* = 1.98, 95% CI BCa [4.23, 5.06]) compared to before (*M* = 6.60, *SD* = 1.31, 95% CI BCa [6.32, 6.87]), *t*_(81)_ = 9.899, *p* <.001, Cohen’s *d* = 1.77.

Among the 2,184 (22.8% of total sample) users who received a recommendation to seek further support in another service outside *krisenchat*, the most frequently recommended services or people for further help seeking included a psychotherapist (*n* = 117, 50.4%), a trusted person (*n* = 27, 11.6%), a counselling service (*n* = 14, 6.0%), and parents (*n* = 13, 5.6%). From those, *n* = 131 (51.1%) followed the recommendation of seeking further help. Missing data was substantial (*n* = 1,916, 87.7%). Of the 131 users who indicated having contacted the recommended service or person, *n* = 85 (64.9%) had already contacted the service and had an appointment, while *n* = 46 (35.1%) had contacted but there was no appointment or they were in the waiting list. Additionally, *n* = 36 (27.5%) participants informed they had a follow-up appointment already scheduled, while *n* = 20 (15.3%) informed they had not follow-up appointment schedule; there was *n* = 75 (57.3%) missing data. No statistical difference by gender was found among those who received a recommendation and reported having sought further help: χ²(2, 266) = 2.717, *p* =.257, ϕc =.101.

Among the 7,398 (77.2%) users who did not receive a recommendation to seek another service, 334 (4.5%) reported having sought further help elsewhere. In this case, there was a statistical difference by gender: χ²(2, 719) = 11.332, *p* =.003, ϕc =.126. Female users (*n* = 286, 48.9%) were more likely to seek help compared to male (*n* = 38, 35.8%) and diverse users (*n* = 7, 25.0%). No significant difference was found between male and diverse users.

The barriers for further help seeking were grouped into stigma (*n* = 172, 46.1%), lack of literacy (*n* = 183, 49.1%), structural (*n* = 60, 16.1%), autonomy (*n* = 129, 34.6%) and family beliefs (*n* = 160, 42.9%). In this case, percentage were calculated based on the total number of cases rather than the number of users; therefore, the sum exceeds 100%. This method was used to identify the most common barriers reported by users.

For all users who reported not seeking further help, regardless of receiving a recommendation, barriers were presented for 373 participants in Table 6. There was no significant difference by gender in barriers to seeking further help (all *p* >.005).

**Table 6.** Barriers to further help seeking for all users who reported not seeking further help, regardless recommendation (*N* = 373).

| Barriers<sup>1,2</sup> | Male | Female | Diverse | Test | p | Effect size |
| --- | --- | --- | --- | --- | --- | --- |
|  | (n = 59) | (n = 297) | (n = 17) |  |  |  |
| | n (%) | n (%) | n (%) | $\chi^2$ | | $\phi_c$ |
| Stigma | 27 (33.3)<sup>a</sup> | 137 (33.3)<sup>a</sup> | 8 (30.8)<sup>a</sup> | (2, 518) = 0.073 | .964 | .012 |
| Lack literacy | 26 (32.1)<sup>a</sup> | 146 (35.5)<sup>a</sup> | 11 (42.3)<sup>a</sup> | (2, 218) = 0.931 | .628 | .042 |
| Structural | 12 (14.8)<sup>a</sup> | 43 (10.5)<sup>a</sup> | 5 (19.2)<sup>a</sup> | (2, 518) = 2.815 | .245 | .074 |
| Autonomy | 20 (33.9)<sup>a</sup> | 100 (33.7)<sup>a</sup> | 9 (52.9)<sup>a</sup> | (2, 518) = 1.385 | .500 | .052 |
| Family beliefs | 23 (39.0)<sup>a</sup> | 132 (44.4)<sup>a</sup> | 5 (29.4)<sup>a</sup> | (2, 518) = 2.182 | .336 | .065 |

Note. <sup>1</sup>Calculation of % from valid cases. <sup>2</sup>Multiple answers were possible. Percentage was calculated based on the total of responses. Cells with the same superscript in a row did not differ significantly.

When examining only those who received a recommendation but did not seek further help (i.e., *n* = 131 users), similar results were observed, with no significant differences by gender in barriers to seek further help (all *p* >.005).

## 4. Discussion

The present study substantially adds to the body of evaluative research that has been undertaken in the past five years on the online crisis chat program *krisenchat* as it is the first analysis to be undertaken with a focus on gender analysis and possible gender differences in utilization, acceptability and help-seeking behavior after service use.

In 2023, the overall uptake of the *krisenchat* service was substantially lower in young males compared to females, which is in line with general access to (online) mental health care offers (29, 30). In comparison to a study conducted on the service in 2021 (13), 4,988 (83.44%) were female, 881 (12.74%) male, and 109 (1.82%) diverse. Further to that, data from the Federal Statistical Office, individuals registered as “diverse” in the 2022 census represented only about 0.001% of the German population and are considered a marginalized group (31).

Male service users were (on average a year) older than female and diverse users. Importantly, young male users in the current sample were significantly less likely to be in previous treatment at the time of contact, stressing the notion that men often seek help later and have fewer prior points of contact with the healthcare system (32). Male *krisenchat* users sent and received fewer messages compared to female and diverse users, albeit with small effect sizes. This finding is in line with research suggesting that males tend to use fewer words in written emotional disclosure, possibly reflecting more ‘task-focused’ communication styles (33, 34). A previous study on *krisenchat* also points towards gender differences in the use of language: female and diverse users used significantly more first-person singular pronouns, insight words, negations, and negative emotion words than male users (16). Men were more likely to access *krisenchat* during late-night and early-morning hours compared to female users and diverse users. Even though *krisenchat* is available as a ‘round-the-clock’ service, this time of day seems to be particularly critical for engaging male users. While specific studies on gender differences in the timing of crisis counselling access are limited, existing research suggests that men may be more likely to seek help during later hours compared to women. For instance, a study found that during the COVID-19 lockdown, men exhibited different patterns in communication and mobility compared to women, which could imply varying help-seeking behaviors at different times of day (35). Additionally, a study by Pretorius et al. (2019) examined online help-seeking behaviors among young people and found that males were more likely to seek help during non-traditional hours, such as evenings, possibly due to factors like work commitments or social expectations (36).

Concerning pathways to become aware (of the service), young males were more likely to discover the service via self-directed search on search engines (Google) whereas young women discovered it through ‘institutional’ (school/university) or social media (TikTok) channels. General preferences of seeking help in young people are as text-based query via an internet search engine. Social media, government or charity websites, live chat, instant messaging, and online communities were also used (36). These findings indicate that tailored approaches to enhance awareness and utilization of services by young males might be necessary.

For general service promotion, this suggests that direct search engine optimization may be especially important for male outreach, whereas integrating services into educational and social platforms may be more effective for female users. Twitch is a platform that has been considered promising for reaching a male target audience. Statistics estimate that 65% of Twitch users are male. Additionally, certain YouTube genres are intensively consumed by boys.

An analysis of topic tags revealed that ‘Emotional distress’ was more frequently tagged as a chat topic by counsellors in young men, potentially functioning as a gendered expression of general psychological strain. This may reflect a linguistic or cultural adaptation in how young men communicate distress in a more indirect way, highlighting the importance of service designs that validate non-suicidal expressions of crisis (16).

Young men were also less likely to report sexual violence or self-harm compared to women, consistent with literature showing male underreporting of victimization and self-injurious behavior due to stigma and gender expectations (37). German data from 2024 indicated a rise of 10% (850 cases) in self-reporting by boys and girls affected by child welfare endangerment (38), indicating there is still room for awareness campaigns through diverse channels.

However, young males were more likely than females to bring up sexuality and LGBTQIA+ topics within *krisenchat* sessions, aligning with findings that anonymous online spaces may provide safer contexts for identity-related disclosure (39).

Interestingly, there was no significant difference between male and female users in the chat topic “suicidality,” nor in suicidal behavior flagged as a chat risk by conselors. In constrast, diverse users were almost twice as likely to report suicidality as a topic and to present suicidal behavior as a chat risk compared to both male and female users. Men are statistically more likely to die by suicide (World Health Organization, 2025), they were generally less likely to disclose suicidal behavior (as tagged by counsellors) compared to diverse users and not significantly different from women. This paradox, also described in prior studies (40, 41), suggests that (young) males’ suicide risk may be underreported in (online) crisis contexts due to barriers in verbalizing suicidality. A scoping review found that women are more likely to contact suicide prevention hotlines than men, despite men having higher suicide rates. This suggests that while women may be more engaged with crisis services, men may be underrepresented in these services, potentially due to factors such as stigma or reluctance to seek help (42).

Despite gender differences in access and utilization, acceptability outcomes across genders were mostly comparable: All genders reported significant reductions in emotional distress post-chat and no differences in satisfaction, self-efficacy, or likelihood of recommending the service. These findings echo meta-analytic evidence suggesting that once engaged, men and women derive comparable therapeutic benefit from online crisis services and telehealth interventions (43, 44). This indicates that digital platforms may reduce some barriers traditionally associated with face-to-face counselling for men (45).

The current study led to similar findings in (15)’s study in terms of following recommendation to seek further support in another (professional) service outside *krisenchat* (48.6% in previous study vs. 51.1% in the current one). Furthermore, no difference by gender was found among those who received a recommendation and reported having sought further help.

Again, regardless of gender, users who did not receive a recommendation to seek another service, a comparable amount (4.5%) reported having sought further help elsewhere. Here, however, female users were more likely to seek help (pro actively/anyways) compared to male and diverse users.

Although no gender differences emerged in uptake following a recommendation, among those not explicitly encouraged, women were more likely to independently seek additional help than men or diverse users. This highlights persistent gender gaps in proactive mental health engagement, echoing population-based findings that women are more likely to initiate formal help-seeking (46).

There was no significant difference by gender in barriers (stigma, low mental health literacy, structural obstacles, desire for autonomy, and family beliefs). A previous study on *krisenchat* (15) and a review including 24 studies (24) suggests that these barriers are largely similar across genders, indicating that while the level of engagement may be lower in males, the types of obstacles faced by males and females are comparable.

### 4.1. Practical Implications

The present results underscore that men are less likely to seek crisis counselling via *krisenchat*, less likely to disclose suicidality while they derive equivalent benefit when they do engage with the service. Digital crisis interventions thus represent a promising way to bridge gender disparities, but their effectiveness in reaching men depends on (a) enhancing discoverability via autonomous search channels/channels preferred by men, (b) tailoring communication strategies to validate diverse expressions of distress, and (c) ensuring follow-up pathways that reduce barriers to sustained engagement in professional care.

### 4.2. Strengths and Limitations

To the best of our knowledge, the present study is the first to conduct a detailed analysis of young male usage of a messenger-based online psychosocial counselling and compare them with female and diverse gender. Some strengths of this study include the recent and large original sample size and the available data during the course of a whole year. Importantly, it highlights that while young men access and use the service differently, they derive comparable benefits, providing actionable implications for tailoring digital mental health interventions.

A limitation concerns the large number of missing data regarding age, as chatters may request deletion of metadata or simply not report their age, leading to the exclusion of many cases. Although the study had an overall large sample size, only 29.6% (*n* = 2,836) of the total sample completed the feedback survey, and 12.4% (*n* = 1,191) completed the follow-up survey.

## 6. Author Contributions

**JH:** Conceptualization, Methodology, Formal analysis, Writing - Original Draft, Writing - Review & Editing. **EK:** Conceptualization, Writing - Original Draft, Writing - Review & Editing. **KJE:** Writing - Original Draft. **ME**: Conceptualization, Investigation, Resources, Writing - Review & Editing. **RW:** Conceptualization, Investigation, Resources, Data Curation, Writing - Review & Editing. **SS:** Conceptualization, Investigation, Data Curation, Writing - Review & Editing. **JP:** Investigation, Writing - Review & Editing. **ASP:** Conceptualization, Methodology, Formal analysis, Writing - Original Draft, Writing - Review & Editing. **CR-K:** Conceptualization, Writing - Review & Editing, Supervision, Project administration. All authors have approved the final manuscript.

## Data Availability

All data produced in the present study are available upon reasonable request to the authors.

